# Abnormal uterine bleeding (AUB) is not a single clinical entity: different clinical causes are associated with distinct cellular and molecular endometrial characteristics

**DOI:** 10.64898/2026.09.14.26360890

**Authors:** Varsha Jain, Florent Petitprez, Jessica Chung, Pia Wahi-Singh, Alistair RW Williams, Aleksandra O Tsolova, Peter AW Rogers, Hilary OD Critchley

## Abstract

**Background:** Abnormal uterine bleeding (AUB) is commonly managed as a single clinical entity with standardised pathways. However, high therapeutic failure and subsequent hysterectomy rates indicate AUB is not a single symptom. AUB affects over a billion individuals worldwide representing a substantial global health burden. While the symptom of AUB has multiple causes, the underlying endometrial mechanisms remain largely unexplored. To determine why standardised, medical management consistently fails, we directly compared the endometrium associated with two distinct aetiologies of AUB: uterine fibroids (AUB-L), and a primary endometrial disorder (AUB-E).

**Methods:** We investigated menstrual cycle-staged endometrium from women with AUB-L (n=32) and AUB-E (n=41). Endometrial tissue underwent quantitative RT-PCR (n=73), bulk RNA sequencing (n=35), and quantitative immunohistochemistry (n=18) to assess transcriptomic signatures and spatial immunolocalisation of sex steroid receptors across menstrual cycle phases.

**Findings:** Deep tissue interrogation confirmed menstrual cycle stage remains the major determinant of gene expression in endometrium of women with AUB and regular menstrual cycles. The global endometrial transcriptome was determined by clinical cause of AUB, with 81 differentially expressed genes (FDR <0·05) identified when comparing AUB-L and AUB-E. Aetiology of AUB dictated endometrial spatial immunolocalisation of sex steroid receptors, with glandular persistence of epithelial progesterone receptor expression (PR, PR-B) in mid-secretory phase endometrium in subjects with AUB-L versus AUB-E.

**Interpretation:** AUB should not be conceptualised as a single endometrial phenotype simply because the clinical bleeding symptom is shared by several aetiologies. These findings demand a paradigm shift in clinical care: to improve patient outcomes, management of AUB must evolve from empirically treating a shared symptom to delivering precision therapeutics guided by distinct molecular endometrial phenotypes.

**Funding:** UK Charity Wellbeing of Women (RTF902), BBSRC-NC3R ageing project grant BB/S002995/1; Wellcome Trust (225021/Z/22/Z); MRC Centre for Reproductive Health Centre grants: G1002033, MR/N022556/1; MRC research grants G0000066, G0500047, G0600048, and MR/J003611/1.

**Research in Context:** *Evidence before this study:* We searched PubMed from inception to July 1, 2026, using terms including “abnormal uterine bleeding”, “endometrium”, “leiomyoma”, and “fibroid” finding a paucity of studies researching the endometrium in women with AUB. Published studies have predominantly focused on the clinical causes of AUB rather than the endometrium, i.e., the tissue that bleeds. Furthermore, when the endometrium has been studied, menstrual cycle phases have frequently been inadequately characterised particularly in the secretory phase, limiting accurate mechanistic inferences. Consequently, the underlying tissue-level mechanistic drivers of AUB remain unknown, and likely contributing to the high failure rates of medical management. It is timely that the biological causes of AUB have been identified recently as the second research priority in the James Lind Alliance Priority Setting Partnership for Problematic Menstrual Bleeding.

*Added value of this study:* We directly compared rigorously phenotyped and menstrual cycle-staged endometrium from women with different clinical causes of AUB (uterine fibroids: AUB-L versus primary endometrial disorder: AUB-E). We demonstrate that the global endometrial transcriptome is determined by the underlying clinical cause of AUB rather than the shared bleeding symptom. Furthermore, we identified that endometrium from women with AUB-L is uniquely characterised by an altered spatial immunolocalisation of sex steroid receptors (PR, PR-B, ERα) during the mid-secretory phase.

*Implications of all the available evidence:* The presence of distinct transcriptomic profiles and altered sex steroid receptor immunolocalisation in the endometrium, provides insights into a biological rationale for why uniform, non-targeted progestin therapies may frequently fail to manage the symptom of AUB. Advancing patient care now necessitates a fundamental conceptual shift. We must cease treating AUB as a singular biological entity, we need to move away from empirical symptom control, and instead move toward precision therapy for AUB guided by exact clinical phenotyping.

## Introduction

Abnormal uterine bleeding (AUB) is a substantial global health burden, affecting up to 50% of women and girls during their reproductive years^1^, which equates to an estimated prevalence of over a billion individuals worldwide^2^. Defined as bleeding from the endometrium that is abnormal in its timing, frequency, or amount^3^, AUB extensively disrupts physical, social, and psychological well-being^4,5^. The socio-economic and healthcare costs are high; in the UK alone, heavy menstrual bleeding (HMB) costs the economy over £5.1 billion annually due to absenteeism and presenteeism^6^. Furthermore, acute-on-chronic HMB requiring emergency blood transfusions for severe iron deficiency anaemia costs the National Health Service (NHS) over £13 million annually^7^. Despite representing one of the most common and debilitating gynaecological presentations globally, AUB remains persistently under-researched^8^. The biological causes of AUB have been identified recently as the second research priority in the James Lind Alliance Priority Setting Partnership for Problematic Menstrual Bleeding^9^.

Under the widely adopted International Federation of Gynecology and Obstetrics (FIGO) AUB system 2 classification^3^, the symptom of AUB is sub-divided by its multiple, diverse underlying causes. Uterine leiomyomas (AUB-L; fibroids) are among the most prevalent structural aetiologies, occurring in 70% of White women and 80% of Black women by age 50^10^. Abnormal bleeding associated with uterine fibroids is typically heavy menstrual bleeding (HMB) which is prolonged and frequent, and is a leading cause of blood transfusion in patients with acute-on-chronic AUB^7^. Conversely, when no specific structural, systemic, or iatrogenic cause is identified following appropriate clinical investigation, the abnormal bleeding is classified as AUB of endometrial origin (AUB-E), presuming the presence of a primary local endometrial disorder. However, it remains unknown whether the endometrium in women with AUB-L shares this same primary endometrial dysfunction, or if structurally distinct aetiologies lead to AUB via alternative molecular mechanisms, i.e. a secondary endometrial disorder^11^.

Despite the profound heterogeneity in aetiology, the medical management of AUB remains empirically uniform^5^. Therapies are currently prescribed using a “one-size-fits-all” approach, treating the symptom of AUB as a single clinical entity with uniform, non-targeted progestins used as first-line medical management^5^. A fundamental barrier to the personalised management of AUB is the current treatment of the symptom as a singular biological entity. Historically, research has compared the endometrium of individuals with AUB to the endometrium of healthy controls. However, understanding why standard therapies fail requires directly comparing the local endometrial mechanisms of distinct aetiologies of AUB, incorporating precise menstrual cycle staging. Recognising this tissue-level heterogeneity is essential to advance patient care and deliver true personalised medicine.

To address this critical knowledge gap, the present study systematically interrogated the transcriptomic and cellular characteristics of the endometrium in well-categorised, menstrual cycle-staged samples from women with AUB associated with uterine fibroids (AUB-L) and AUB associated with a primary endometrial disorder (AUB-E). By diagnosing AUB-E through the strict clinical exclusion of all other causes of AUB as per the FIGO AUB System 2 classification recommendations^3^, we utilised AUB-E as a clinically defined comparator to evaluate the abnormal bleeding endometrium without the influence of a structural cause. We present novel molecular data demonstrating that AUB must no longer be conceptualised as a single endometrial phenotype merely because the outward clinical symptom is shared. We establish that clinical aetiology of AUB alters the endometrial transcriptome and reveals an altered spatial immunolocalisation of sex steroid receptors, particularly in the mid-secretory phase. Deep molecular interrogation confirms that clinical causes of AUB influence endometrial biology, demanding a shift away from empirical treatment and toward precision phenotyping in clinical gynaecology.

## Methods

### Study design, ethics and human patient cohort

Endometrial samples were obtained from a reproductive tract tissue resource archive in which each sample is linked to clinical phenotyping metadata from the originating patient. Ethical approvals for the use of all endometrial samples were granted by NHS Health Research Authority’s Research Ethics Committees (Refs. 19/SS/0102 and 20/ES/0119), and all participants provided written informed consent. Patients were recruited after attending gynaecology services within NHS Lothian hospitals and surplus endometrial tissue from clinically indicated biopsies or hysterectomies (performed for the primary symptom of AUB) were used for research purposes.

### Clinical phenotyping and inclusion criteria

Samples were selected from reproductive-aged women (aged 26 - 52 years) presenting with the primary complaint of either AUB-L or AUB-E (Supplementary Table 1). Criteria for inclusion and exclusion are detailed in Table 1:

**Table 1:** Inclusion and exclusion for participation in this current study.

| Inclusion Criteria | Exclusion Criteria |
| --- | --- |
| Main clinical complaint of AUB (predominantly heavy menstrual bleeding, HMB) | Use of exogenous hormones within two months prior to endometrium sampling |
| Regular menstrual cycles of 21-35 days | Current use or use within two months of selective progesterone receptor modulators (SPRMs), gonadotrophin releasing hormone (GnRH) analogues or GnRH receptor antagonists, |
| Endometrial samples consistent across stage of menstrual cycle (proliferative, early secretory, early-mid secretory, mid-secretory) when assessed using three parameters:<br>(a) standard histological criteria<sup>12</sup>,<br>(b) serum oestradiol and serum progesterone level at the time of endometrium sampling, and<br>(c) clinical menstrual cycle history. | Current steroid use, |
|  | Evidence or suggestion of endometriosis, |
|  | Previous endometrial ablation or uterine artery embolisation, |
|  | Any other cause of AUB clinically identified using the FIGO AUB System 2 classification<sup>3</sup> i.e., Polyps, Adenomyosis, Endometrial hyperplasia/Endometrial Cancer, Coagulopathy, Ovulatory Dysfunction, Iatrogenic, Not otherwise classified. |

For participants with AUB-L, endometrial tissue was derived from those with imaging (ultrasound or MRI) or histopathological evidence of intramural fibroids (at least one fibroid, >3cm), i.e., fibroid within the uterus surrounded entirely by myometrium, and not in contact with the endometrium or the serosa of the uterus, i.e., intramural fibroids - FIGO types 3 and 4^3^. Endometrial tissues derived from women with submucous fibroids or only subserosal fibroids were excluded to ensure observed endometrial changes were not physical effects of uterine cavity distortion.

### Primary menstrual cycle staging

Endometrial samples were allocated to distinct menstrual cycle stages (proliferative, early secretory, early-mid secretory, mid-secretory) utilising three concurrent parameters: (i) standard histological criteria^12^, (ii) serum oestradiol and serum progesterone level measured at the time of endometrium sampling, and (iii) clinical menstrual cycle history. These three criteria are collectively hereafter referred to as the “Edinburgh classification” of the endometrium. Late secretory and menstrual phase samples were excluded as these stages are characterised by a decline in circulating progesterone concentrations due to luteal regression.

A total of 73 samples met inclusion criteria and were interrogated using quantitative reverse transcriptase polymerase chain reaction (RT-qPCR, n=73). A cohort of samples were examined with bulk RNA sequencing (n=35), and another cohort of samples were investigated using immunohistochemistry (n=18) (Supplementary Table 1).

### Quantitative Reverse Transcriptase Polymerase Chain Reaction (RT-qPCR)

Total RNA was extracted from endometrial samples using the Qiagen RNeasy mini kit as per the manufacturer’s guidance (Qiagen, UK). RNA concentration was estimated using Nanodrop spectrophotometer (ThermoFisher Scientific, UK). Complementary DNA (cDNA) was synthesised using the iScript cDNA synthesis kit (Bio-Rad Laboratories, UK) with a 100ng RNA template. RT-qPCR was conducted in triplicate reactions using TaqMan primers and probes (Roche Diagnostics, UK) for a group of candidate genes including sex steroid receptors: progesterone receptor (*PGR*), oestrogen receptor (*ESR1),* and candidate progesterone-dependent genes: Interleukin 15 (*IL15*), Heart- and Neural Crest Derivatives-Expressed Protein 2 (*HAND2*), Forkhead Box O1 (*FOXO1*), Homeobox A10 (*HOXA10*), 17β-Hydroxysteroid dehydrogenase 2 (*HSD17B2*). All candidate genes selected are known to have expression which is dependent on progesterone^13–19^. The primers and probes were validated for efficiency prior to use (Supplementary Table 2). Target gene mRNA expression levels were normalised to the geometric mean of the endogenous reference genes ATP synthase H transporting mitochondrial F1 complex beta polypeptide (*ATP5F1B*) and succinate dehydrogenase (*SDHA*). Relative expression was calculated against a calibrator endometrial sample (mixture of proliferative and secretory endometrium) using the comparative ΔΔCT method^20^.

### RT-qPCR analysis

Statistical analyses of mRNA expression were performed using GraphPad Prism version 10·0 (San Diego, CA). RT-qPCR data are presented as scatter dot plots with median, maximum and minimum points demonstrated. Data were analysed to understand the effect of menstrual cycle stage as determined by the “Edinburgh classification” of the endometrium (as described above), and non-parametric Kruskal-Wallis test was used to compare all menstrual cycle stages against each other. For an overall effect of menstrual cycle stage, all data from AUB-E and AUB-L samples were pooled together. The effect of clinical cause of AUB was analysed by separating AUB-E and AUB-L per menstrual cycle stage and comparing data for each menstrual cycle stage directly, using non-parametric Mann-Whitney U test. Statistical significance was defined as *p* <0·05 for all analyses.

### Bulk RNA sequencing analysis

A cohort of extracted RNA from endometrial samples within this study (Proliferative: AUB-L n=6, AUB-E n=6; early secretory: AUB-L n=5, AUB-E n=2; early-mid secretory: AUB-L n=1, AUB-E n=4, mid-secretory: AUB-L n=5, AUB-E n=6), were interrogated using bulk RNA sequencing using RNA sequencing platform NextSeq 550 (Illumina Inc, #SY-415-1002) in two separate batches. Library preparation utilised the QuantSeq 3’ mRNA-Seq Library Prep Kit (FWD) for Illumina (Lexogen Inc, #015). Raw reads were pre-processed into gene counts using Lexogen Kangooroo BETA (Lexogen GmBH, Austria). Menstrual cycle stage was classified using the “Edinburgh classification” of the endometrium (as described above).

### Molecular Menstrual Cycle Timepoint Classification

To determine menstrual cycle timepoint continuously for further analyses, bulk RNA sequencing transcriptomic data were processed using Endest version 0·1·1^21^. This statistical algorithm utilises the entire transcriptome to assign a continuous numerical timepoint from 0% to 100% across the entire menstrual cycle, representing the menstrual (0-8%), proliferative (8-58%), early secretory (58% - 72%), mid-secretory (72% - 87%), and late secretory (87% - 100%) phases^21^. This menstrual cycle timepoint is independent of categorical staging which occurred as per the “Edinburgh classification” of the endometrium (Supplementary Table 1).

### Differential gene expression and pathway analysis

Lowly expressed genes were removed by requiring > 0·5 counts per million (CPM) in > 50% of samples. Counts were normalised for library size and composition bias using the Trimmed Mean of M-values (TMM) method^22^. Normalised counts were transformed into log2 CPM using voom^23^, and a linear model was fitted to each gene using limma v3·60·2^24^ in the R programming language.

To determine differences associated with the clinical causes of AUB, while controlling for cycle variance, all linear models included the clinical cause of AUB (AUB-L versus AUB-E), sequencing batch, and the Endest^21^-derived continuous menstrual cycle timepoint. The menstrual cycle timepoint was modelled as a natural cubic spline with 8 degrees of freedom. Empirical Bayes moderated t-statistics assessed differential expression, with multiple hypothesis correction via the Benjamini-Hochberg method. Genes were considered significant if the adjusted p-values, i.e., the false discovery rate (FDR) was < 0·05. Gene ontology analyses were performed with clusterProfiler v4·12·2^25^ using genes with a significance level of FDR < 0·1.

### Immunohistochemistry

A cohort of endometrial tissue (formalin-fixed paraffin embedded) samples from the proliferative (AUB-E n=5, AUB-L n=6) and mid-secretory (AUB-E n=4 AUB-L n=3) phase were studied for expression patterns of progesterone receptor (PR), progesterone receptor B (PR-B), and oestrogen receptor alpha (ERα) (Supplementary Table 3). Immunohistochemistry (IHC) was performed using the Leica Bond III Stainer (Leica Microsystems, IL, USA). Paraffin sections were dewaxed, rehydrated, and processed using the Leica Bond Polymer Refine Detection Kit (DS9800, Leica Microsystems) with 3,3′-diaminobenzidine (DAB) visualization. Antigen retrieval was carried out using Bond Epitope Retrieval Solution 1 (ER1; pre-diluted, pH 6·0; AR9961). Endogenous peroxidase activity was blocked with 3% hydrogen peroxide, and slides were washed in BOND wash (TBS-T). Antibody-specific protocols were applied according to host species, with all antibodies optimised for dilution prior to analysis (supplementary table 3). Slides were counterstained, dehydrated, mounted, and whole slides were scanned using an Axioscan 7 slide scanner for digital image analysis. Slide images were reviewed using the Zen Blue (Carl Zeiss Microscopy GmbH, version 3·7·97·00000) and representative photomicrographs were captured.

### Digital Image Analysis

Digital image analysis of immunohistochemistry studies was performed, blind to clinical phenotype and histological sample categorisation, using QuPath (version 0·5·1-x64) to quantify positive staining of PR, PR-B, and ERα nuclear markers in the stromal and glandular epithelial compartments of the endometrial samples^26^. Analysis settings were optimised for sex steroid hormone receptor staining which was conducted using DAB as above. A whole-slide annotation was created, excluding areas of luminal epithelium (not all samples contained luminal epithelium), areas in which the tissue architecture was lost, or areas with staining discrepancies, so that the analyses focused only on functional endometrium, i.e., upper layer of the endometrium closest to the luminal epithelium. Cells were detected using Qupath’s Cell Detection function. A cell classifier was trained to identify glandular epithelial cells and stromal cells. The positive DAB staining threshold was set to identify DAB positive, and DAB negative cells and the percentage of positive stained cells were further used for comparative analyses. Non-parametric Mann-Whitney U test was used to compare AUB-L and AUB-E staining of the sex steroid hormone receptors per menstrual cycle stage. Statistical significance was defined as *p* <0·05.

## Results

### Clinical phenotyping defines distinct underlying causes of AUB

To separate the shared symptom of bleeding from underlying molecular pathology, we established a novel framework to systematically interrogate the endometrium using both targeted and unsupervised molecular measures (Figure 1A). First, endometrial samples from women presenting with abnormal uterine bleeding (AUB) were first subjected to clinical phenotyping. Patients were stratified into clinically defined cohorts: AUB associated with uterine fibroids (AUB-L), with clinical confirmation that the fibroids did not distort the endometrial cavity, and AUB associated with a primary endometrial disorder (AUB-E) (Figure 1A). A comparison of clinical characteristics (Table 2) revealed that women with AUB-L were older than those with AUB-E (45 vs. 41 years median age in the proliferative stage, p = 0·0086; 44 vs. 32 years in the early secretory stage, p=0·026, 45 vs. 41 years median age in the mid-secretory stage, p = 0·044). BMI, parity, clinical menstrual cycle history, and serum sex steroid hormone levels on the day of sampling were matched between groups, isolating clinical aetiology as the primary variable.

**Figure 1:**
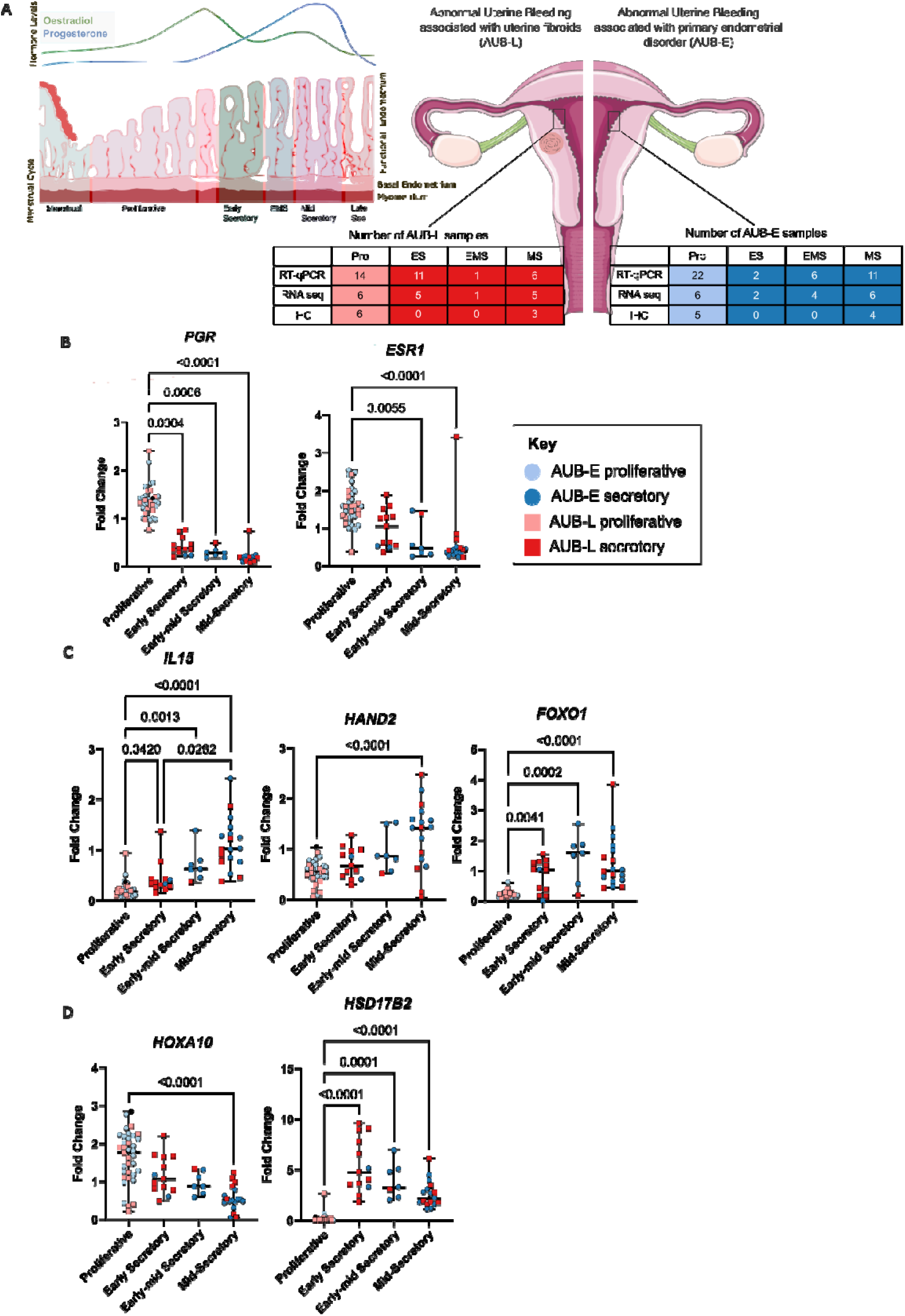
Menstrual cycle stage is the major functional determinant influencing cycling endometrium from women with AUB. 1A – The endometrium adapts across the menstrual cycle in response to changing levels of systemic sex steroid hormones: oestradiol and progesterone with corresponding menstrual cycle phases: menstrual, proliferative (pro), early secretory (ES), early-mid secretory (EMS), mid secretory (MS), and late secretory (late sec). Endometrial samples from women with AUB associated with uterine fibroids (AUB-L) or a primary endometrial disorder (AUB-E) from the pro, ES, EMS, and MS were categorised by menstrual cycle stage (classification as detailed in Methods) and investigated using RT-qPCR (n=73), bulk RNA sequencing (RNA seq) and immunohistochemistry (IHC). 1B – mRNA expression of sex steroid receptors progesterone receptor (PGR) and oestrogen receptor (ESR1) across the menstrual cycle using RT-qPCR. 1C – mRNA expression of progesterone dependent genes: Interleukin 15 (IL15), Heart- and Neural Crest Derivatives-Expressed Protein 2 (HAND2), Forkhead Box O1 (FOXO1), across the menstrual cycle. 1D – mRNA expression of progesterone dependent genes: Homeobox A10 (HOXA10), 17β-Hydroxysteroid dehydrogenase 2 (HSD17B2), across the menstrual cycle, using RT-qPCR. In figures 1B-D: AUB-E are depicted as blue circles (proliferative phase - light blue, secretory phases - dark blue); AUB-L are depicted as red squares (proliferative phase - light red, secretory phase - dark red). Statistical test: Kruskal-Wallis test. Only significant p-values <0·05 have been marked. Figure created by authors using a licensed version of GraphPad Prism, open source Inkscape software and the image of uterus in panel 1A was provided by Servier Medical Art (https://smart.servier.com), licensed under CC BY 4.0. https://creativecommons.org/licenses/by/4.0/).

**Table 2:** Clinical characteristics, clinical history and serum oestradiol and progesterone levels on the day of endometrial sampling, sub-divided into menstrual cycle stages as per the “Edinburgh classification” of the endometrium. Menstrual cycle timepoints (%) provided as calculated by the Endest^21^ algorithm. Data provided as median with interquartile range. There was only one endometrial sample allocated to the early-mid secretory stage from women with AUB associated with uterine fibroids (AUB-L) therefore interquartile range omitted and Mann Whitney test not possible.

|  | Endometrium from women with AUB associated with a primary endometrial disorder (AUB-E) | Endometrium from women with AUB associated with uterine fibroids (AUB-L) | Mann Whitney test *p*-value |
| --- | --- | --- | --- |
| **Age in years** |  |  |  |
| Proliferative | 41 (36-44) | 45 (43-48) | **0.0086\*** |
| Early Secretory | 32 (31-32) | 44 (41-46) | **0.026\*** |
| Early-mid Secretory | 40 (35-42) | 47 | N/A |
| Mid-secretory | 41 (39-43) | 45 (43-48) | **0.044\*** |
| **BMI kg/m<sup>2</sup>** |  |  |  |
| Proliferative | 29 (21-34) | 29 (23-36) | 0.88 |
| Early Secretory | 22 (22) | 26 (22-30) | N/A |
| Early-mid Secretory | 33 (32-38) | 24 | N/A |
| Mid-secretory | 25 (23-28) | 27 (22-27) | 0.99 |
| **Parity** |  |  |  |
| Proliferative | 2 (1-3) | 2 (1-2) | 0.24 |
| Early Secretory | 2 (2) | 3 (2-3) | 0.67 |
| Early-mid Secretory | 2 (2-2) | 2 | N/A |
| Mid-secretory | 1 (0-2) | 2 (1-2) | 0.32 |
| **Menstrual Cycle Length** |  |  |  |
| Proliferative | 28 (25-30) | 28 (28-30) | 0.71 |
| Early Secretory | 29 (28-29) | 28 (28-30) | 0.98 |
| Early-mid Secretory | 28 (28-28) | 28 | N/A |
| Mid-secretory | 29 (28-30) | 28 (27-30) | 0.38 |
| **Menstrual Cycle Day** |  |  |  |
| Proliferative | 11 (10-15) | 11.5 (8-14) | 0.58 |
| Early Secretory | 19 (18-21) | 18 (16-21) | >0.99 |
| Early-mid Secretory | 18 (15-18) | 18 | N/A |
| Mid-secretory | 24 (23-26) | 23 (15-24) | 0.44 |
| **Serum Oestradiol (pmol/l)** |  |  |  |
| Proliferative | 298 (176-579) | 459 (244-827) | 0.31 |
| Early Secretory | 483 (445-522) | 458 (286-651) | >0.99 |
| Early-mid Secretory | 423 (280-464) | 321 | N/A |
| Mid-secretory | 342 (285-440) | 442 (370-587) | 0.31 |
| **Serum Progesterone (nmol/l)** |  |  |  |
| Proliferative | 1.1 (0.6-2.4) | 2.6 (0.7-4.0) | 0.45 |
| Early Secretory | 50 (45-55) | 42 (27-57) | 0.77 |
| Early-mid Secretory | 44 (28-26) | 22 | N/A |
| Mid-secretory | 38 (24-73) | 31 (25-35) | 0.43 |
| **Menstrual cycle timepoint using Endest<sup>21</sup> algorithm (%)** |  |  |  |
| Proliferative | 27 (24-28) | 34 (22-45) | 0.78 |
| Early Secretory | 66 (66-67) | 65 (63-65) | 0.52 |
| Early-mid Secretory | 71 (63-72) | 60 | N/A |
| Mid-secretory | 79 (77-80) | 76 (73-79) | 0.50 |

Crucially, to ensure that menstrual cycle phase did not confound our downstream molecular analyses, we accounted for cycle stage using stringent multi-parameter evaluation. Endometrial samples (n=73) were categorised as proliferative, early secretory, early-mid secretory, and mid-secretory, using the “Edinburgh classification” of the endometrium, a combined assessment of three parameters: (i) histological classification^12^, (ii) circulating serum oestradiol and progesterone levels at the same time of endometrial sampling, and (iii) detailed clinical menstrual cycle history. Furthermore, we used this biological staging to validate the transcriptomic Endest^21^ algorithm (Table 2). This confirmed that computationally generated continuous menstrual cycle timepoints aligned with our assignments of menstrual cycle phase and did not differ significantly between AUB-L and AUB-E samples. This extensive stratification for both clinical aetiology and menstrual cycle stage provided a robust framework to systematically interrogate the endometrium using targeted and unsupervised molecular measures.

### Menstrual cycle stage dictates endometrial gene expression in women with AUB

To determine the baseline influence of menstrual cycle stage on sex steroid receptors and candidate progesterone-dependent genes, a supervised approach was applied. All endometrial samples were initially pooled and analysed via RT-qPCR.

RT-qPCR revealed that the median mRNA expression of sex steroid receptors (*PGR, ESR1*) was highly dependent on the specific phase of the menstrual cycle (Figure 1B). Both receptors were expressed at their highest levels in the proliferative phase, followed by a relative decrease across the secretory continuum. Progesterone receptor (*PGR*) showed significantly higher mRNA expression in proliferative phase samples compared to either early secretory, early-mid secretory, or mid-secretory samples (Figure 1B). *ESR1* expression was significantly higher in proliferative phase samples compared to early-mid secretory or mid-secretory phase samples (Figure 1B). There were no other significant comparisons.

Similarly, menstrual cycle stage significantly influenced the mRNA expression of all progesterone-dependent genes assessed (*IL15, HAND2, FOXO1, HOXA10, HSD17B2*) (Figures 1C and 1D). *IL15, HAND2*, and *FOXO1* mRNA expression levels (Figure 1C) were relatively lowest in the proliferative phase, increasing across the secretory phase from early-secretory to early-mid secretory to mid-secretory phases. *IL15* and *FOXO1* mRNA expression was significantly lower in the proliferative compared to early-secretory, early-mid secretory, and mid-secretory phases. *HAND2* mRNA expression was significantly lower in the proliferative versus mid-secretory phase only. Within the secretory phase, *IL15* demonstrated a significantly lower mRNA expression in the early-secretory compared to the mid-secretory phase.

In this assessment, *HOXA10* (Figure 1D) showed relatively higher endometrial expression in the proliferative phase, followed by a relative decrease across the secretory phase from early-secretory to early-mid secretory to mid-secretory phases. There was significance when comparing the higher proliferative phase *HOXA10* mRNA expression with the lower mid-secretory phase mRNA expression. *HSD17B2* mRNA expression was relatively lowest in the proliferative phase, significantly peaked in the early secretory phase, and subsequently reduced in the early-mid secretory and mid-secretory phases. There was significance when comparing the lower proliferative phase *HSD17B2* mRNA with the early-secretory, early-mid secretory, and mid-secretory phases.

Overall, every candidate gene (*PGR, ESR1, IL15, HAND2, FOXO1, HOXA10, HSD17B2*) demonstrated significant differences between the proliferative versus mid-secretory phase. This confirms that precise determination of menstrual cycle stage is a critical prerequisite for identifying underlying cellular and molecular pathologies in the endometrium, regardless of the clinical cause of AUB.

### Endometrial expression of targeted genes does not distinguish aetiology of AUB

Having established the profound influence of menstrual cycle staging in the endometrium from women with AUB, we next evaluated whether the clinical cause of AUB (AUB-L versus AUB-E) altered the endometrial expression of candidate genes within each specific phase (Figure 2). There was only one endometrial sample classified as being in the early-mid secretory phase from a participant with AUB-L; therefore, pairwise comparison in this specific phase could not be conducted.

**Figure 2:**
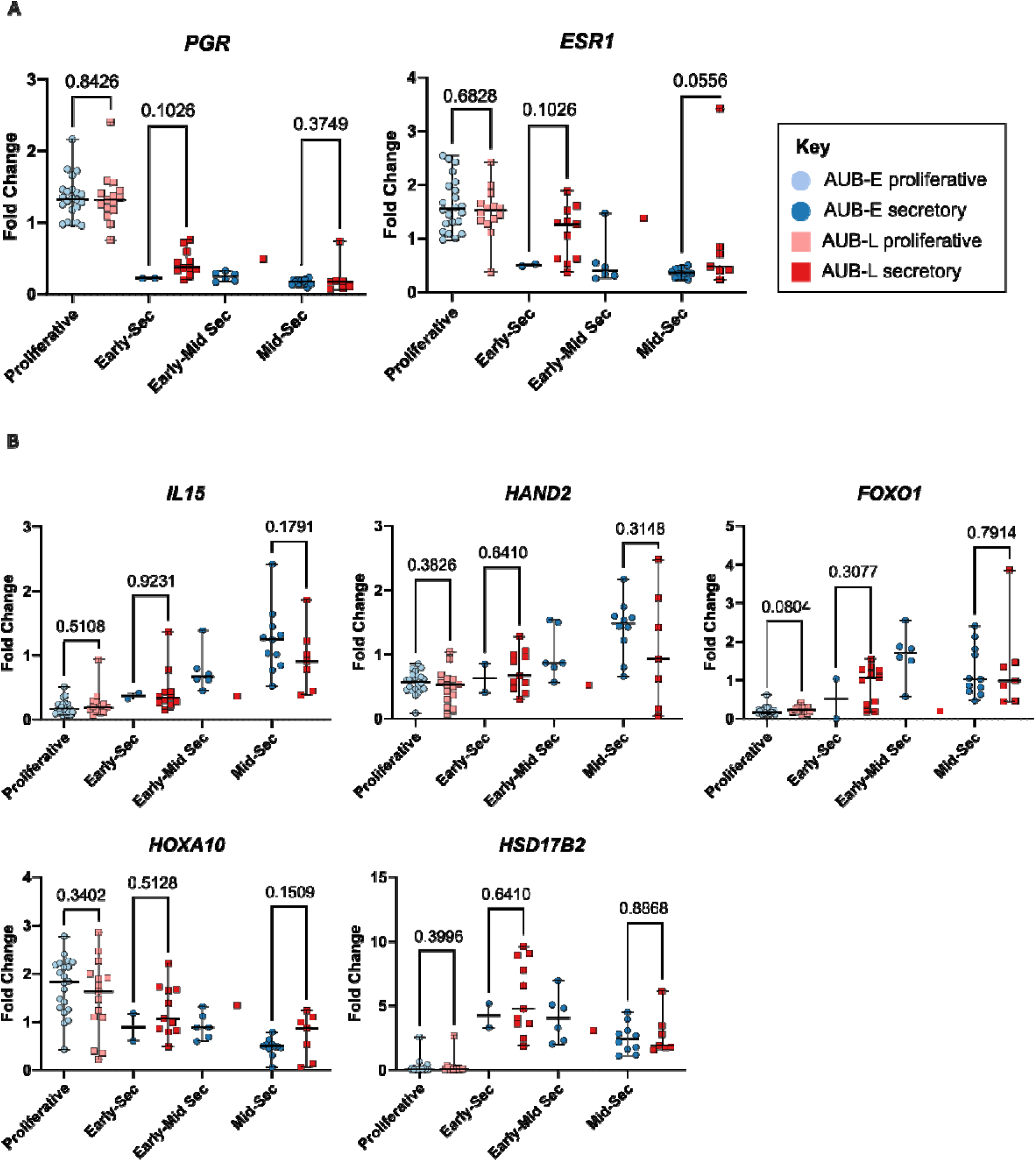
Comparing clinical cause of AUB does not reveal differences in endometrial sex steroid receptor or candidate progesterone dependent genes. 2A – endometrial mRNA expression of sex steroid receptors progesterone receptor (PGR) and oestrogen receptor (ESR1) across the menstrual cycle with pair-wise comparisons made between the expression in endometrium from women with abnormal uterine bleeding associated with uterine fibroids (AUB-L, red squares) and endometrium from women with a primary endometrial disorder (AUB-E, blue circles). 2B – mRNA expression of progesterone dependent candidates: Interleukin 15 (IL15), Heart- and Neural Crest Derivatives-Expressed Protein 2 (HAND2), Forkhead Box O1 (FOXO1), Homeobox A10 (HOXA10), 17β-Hydroxysteroid dehydrogenase 2 (HSD17B2), across the menstrual cycle, with pair-wise comparisons made (statistical test: Mann-Whitney U test) between AUB-L and AUB-E. Proliferative phase samples are in a lighter colour, secretory phase samples are depicted in a darker colour. All p-values have been marked. Figure created by authors with a licensed version of Graphpad Prism and open source Inkscape software.

All pair-wise comparisons between AUB-L and AUB-E from matched cycle stages (proliferative, early secretory, and mid-secretory) revealed no significant differences in the mRNA expression of sex steroid receptors (*PGR, ESR1*) (Figure 2A) or progesterone-dependent genes (*IL15, HAND2, FOXO1, HOXA10, HSD17B2*) (Figure 2B). Therefore, when utilising a targeted, bulk-tissue RT-qPCR approach, distinct clinical causes of AUB did not exhibit significantly different mRNA expression profiles.

### The global endometrial transcriptome is determined by the underlying clinical cause of AUB

To determine whether this apparent targeted candidate approach masked broader molecular differences, an unsupervised approach via whole-transcriptome RNA-sequencing was employed for 35 samples (AUB-L, n=17; AUB-E, n=18). Endometrial samples classified as proliferative, early-secretory, and mid-secretory phases using the “Edinburgh classification” of the endometrium (as defined in Methods), were studied.

Principal component analyses (PCA) provided a global overview of data variability (Figure 3A). While overall gene expression appeared relatively homogenous for proliferative phase samples regardless of the aetiology of AUB, distinct transcriptomic heterogeneity emerged during the secretory phases between the two clinical groups. The Endest^21^ algorithm was used to assign a numerical value between 0% and 100% to represent the exact molecular menstrual cycle timepoint as a continuum where 0% would represent the start of menstruation and 100% would represent the end of the late secretory phase. The molecular menstrual cycle staging timepoint was determined by the Endest^21^ algorithm and was found to align with the “Edinburgh classification” of the endometrium menstrual cycle stage (Figure 3B). Importantly, continuous menstrual cycle timepoints derived from the algorithm did not differ significantly between AUB-L and AUB-E samples within their assigned stages. (Table 2) Endometrial samples studied herein had menstrual cycle timepoints ranging from 11%-84%^21^ (Supplementary Table 1), which corresponds to the proliferative to mid-secretory phases as determined by the histological assessment of the endometrium^21^, (see Methods).

**Figure 3:**
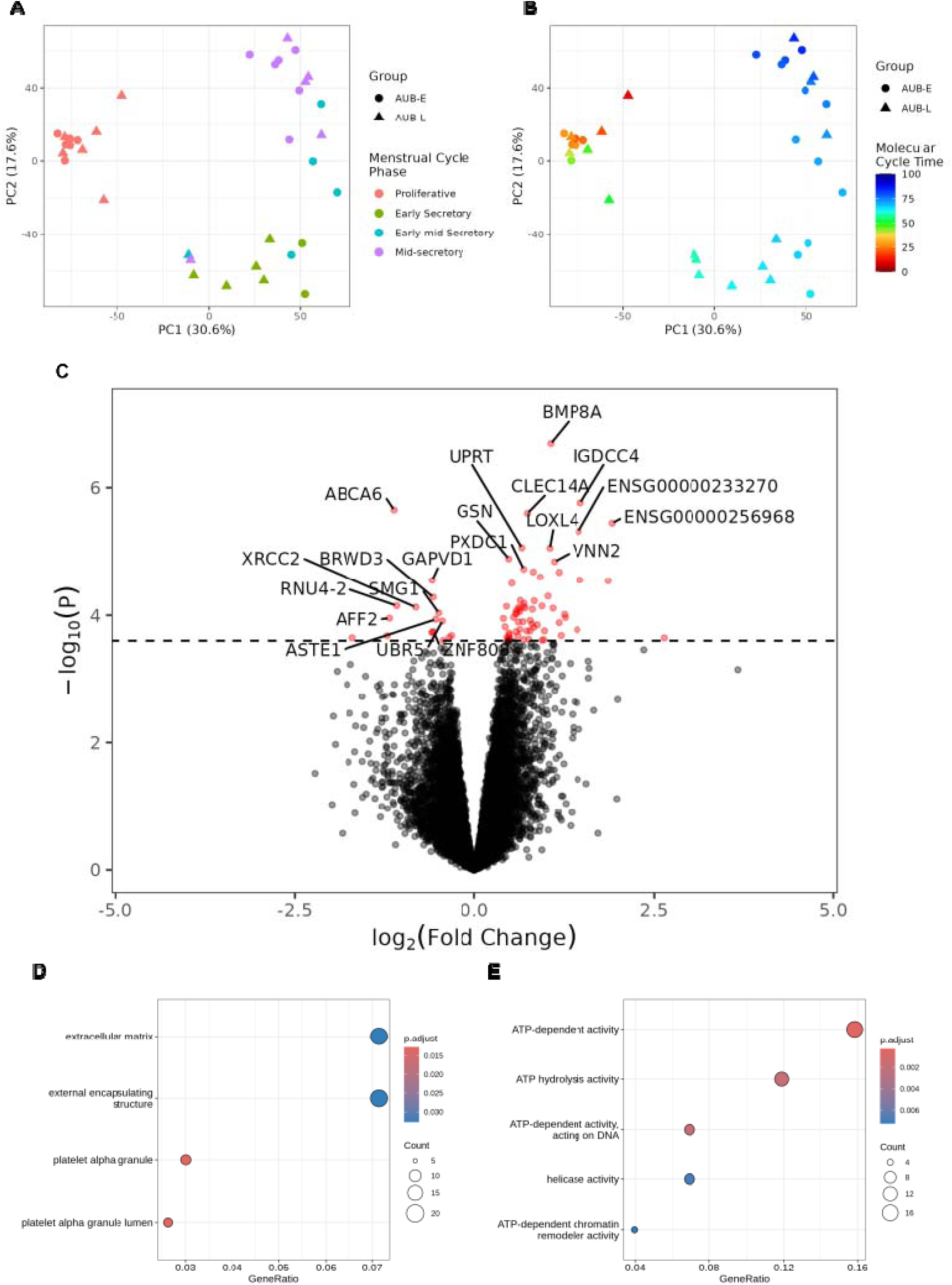
The endometrial transcriptome is strongly influenced by menstrual cycle stage and is distinct in the presence of uterine fibroids. 3A – Principal component plots for first and second components from principal component analyses (PCA) using data derived from RNA sequencing conducted on endometrium from women with AUB-L (n=17) or endometrium from women with AUB-E (n=18), either in the proliferative, early secretory, early-mid secretory or mid-secretory phases – as defined in Methods. The PCA plot shows a characteristic cyclic pattern with samples aligning to menstrual cycle stage order. 3B – PCA plot with samples coloured using menstrual cycle time determined using the Endest^21^ algorithm and alignment with menstrual cycle timepoints ranging from 11%-84% (where 0% represents the start of the menstrual phase and 100% represents the end of the late secretory phase). 3C – volcano plot depicting the differentially expressed gene (DEG) comparison of the endometrial transcriptome between AUB-L vs AUB-E. The y-axis corresponds to the p-value before correction for multiple testing and the horizontal dashed line corresponds to FDR = 0·05. Each point represents a gene, plotted by log2 fold change and -log10 p-value. 81 significantly DEGs (FDR < 0·05) are highlighted in red, with DEGs significantly downregulated in AUB-L on the left of the plot with negative log_2_ fold change, and DEGs significantly upregulated in AUB-L on the right of the plot, with positive log_2_ fold change. Top 10 significant genes in each direction are labelled. 3D and 3E - Dotplots to demonstrate the gene ontology (GO) terms identified to be significantly over-represented when conducting a gene ontology enrichment analysis using the 81 significant DEGs. GO terms have been divided as those which were identified to be upregulated (3D) or downregulated (3E), in the endometrium from women with AUB-L when compared to endometrium from women with AUB-E.

Differential gene expression analyses compared the entire endometrial transcriptome from AUB-L versus AUB-E, with menstrual cycle timepoint (determined by Endest^21^) modelled as a spline to account for precise menstrual cycle staging for all samples analysed. This identified 81 differentially expressed genes (DEGs) (Figure 3C, Supplementary Table 4). Of these, 64 DEGs were significantly upregulated, whilst 17 DEGs were significantly downregulated in AUB-L compared to AUB-E.

Gene Ontology enrichment analyses were conducted on these significantly regulated DEGs to map discrete biological processes (Figure 3D and 3E, Supplementary Table 5). Pathways related to platelet alpha granules, extracellular matrix, and external encapsulating structures were significantly over-represented among upregulated genes in AUB-L (adjusted p<0·05). Conversely, pathways related to ATP-dependent activity, ATP hydrolysis, helicase activity, and ATP-dependent chromatin remodelling were significantly downregulated in AUB-L compared to AUB-E (adjusted p<0·05). Overall, with adjustment for precise menstrual cycle stage timing, unbiased global transcriptomic profiling of the endometrium reveals that distinct biological pathways are altered in AUB-L compared to AUB-E.

### Spatiotemporal localisation of sex steroid receptors reveals distinct endometrial phenotypes differentiated by clinical cause of AUB

As bulk transcriptomic analyses average gene expression across a multicellular tissue and potentially obscure critical spatial dynamics, quantitative immunohistochemical analysis was undertaken at the protein level. To assess spatial dynamics potentially obscured by transcriptomic analyses (Figures 1B, 1C, 3C), we quantitatively mapped the compartment-specific spatial localisation of progesterone receptor (PR), progesterone receptor B (PR-B), and oestrogen receptor alpha (ERα) using QuPath^26^ (AUB-L: n=6 proliferative phase, n=3 mid-secretory phase; AUB-E: n=5 proliferative phase, n=4 mid-secretory phase).

In the proliferative phase (Figure 4A), the spatial immunolocalisation of PR and PR-B in both stromal and glandular epithelial cells was comparable between AUB-L and AUB-E (Figure 4B). However, ERα expression was already significantly lower in the endometrial stromal cells of AUB-L compared to AUB-E (Figure 4B). Spatial immunolocalisation of ERα was similar in the proliferative phase glandular epithelial cells in AUB-E and AUB-L.

**Figure 4:**
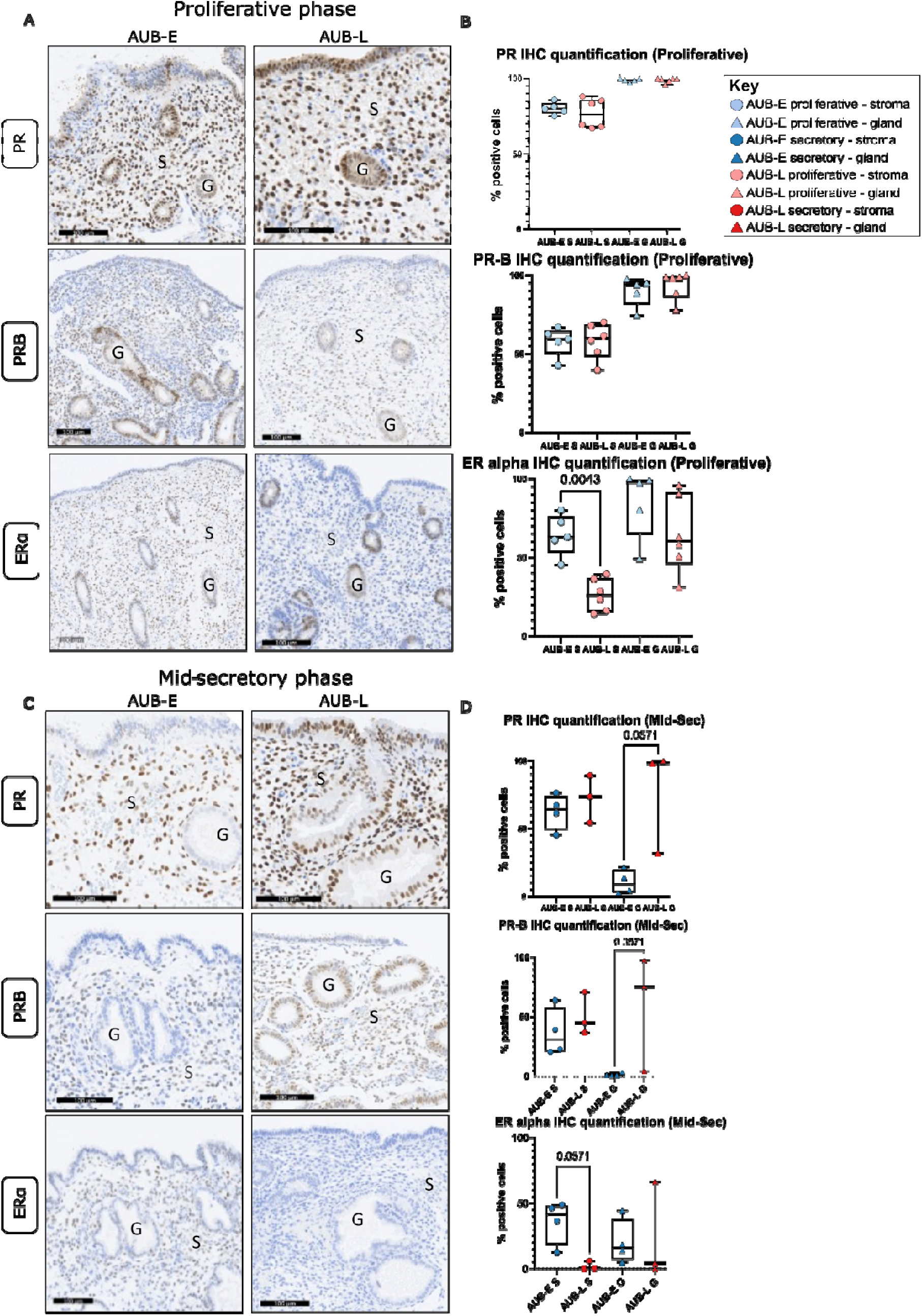
Spatial immunolocalisation of progesterone and oestrogen receptors differ according to clinical cause of AUB. 4A – representative images of immunohistochemical staining (IHC) of progesterone receptor (PR), progesterone receptor B (PR-B), and oestrogen receptor (ERα) in the proliferative phase endometrium of women with either AUB associated with fibroids (AUB-L) or a primary endometrial disorder (AUB-E). Positive immunostaining is brown, negative immunostaining is blue. Glandular epithelium (G) and stroma (S). Scale bars represent 100μm. 4B – Quantitative analyses of the positive immunostaining of PR, PR-B, and ERα, in proliferative phase AUB-E and AUB-L in the stromal (circles) and glandular epithelium (triangles), with quantification conducted using QuPath^26^. Statistical test: Mann-Whitney, significance set at p<0·05. 4C – representative images of immunohistochemical staining of PR, PR-B and ERα, in mid-secretory (mid-sec) phase AUB-E or AUB-L. Positive immunostaining is brown, negative immunostaining is blue. Glandular epithelium (G) and stroma (S). Scale bars represent 100μm. 4D – Quantitative analyses of the positive immunostaining of PR, PR-B, and ERα, in mid-secretory phase AUB-E and AUB-L in the stromal (circles) and glandular epithelium (triangles), with quantification conducted using QuPath^26^. Statistical test: Mann-Whitney U test, significance set at p<0·05. Figure created by authors with a licensed version of Graphpad Prism and open source Inkscape software.

Importantly, greater spatial difference emerged during the mid-secretory phase (Figure 4C). AUB-L featured increased expression of both PR and PR-B in the glandular epithelium, which showed near significance when compared to AUB-E. AUB-E endometrium demonstrated a virtual complete loss of epithelial PR and PR-B expression. The expression of both PR and PR-B was found at comparable levels in stromal cells in mid-secretory phase endometrium (Figure 4D). Furthermore, there was an almost complete loss of ERα expression in the endometrial stromal cells of AUB-L compared to AUB-E, which approached significance. Endometrial glandular epithelial expression of ERα remained similar between AUB-L and AUB-E.

Taken together, these data establish that AUB-L and AUB-E exhibit distinctly different compartment-specific localisations of sex steroid receptors. Specifically, AUB-L is characterised by greater glandular epithelial immunolocalisation of PR and PR-B in the mid-secretory phase, accompanied by reduced stromal cell immunolocalisation of ERα across both the proliferative and mid-secretory phases.

## Discussion

This study establishes that the endometrium from women with abnormal uterine bleeding (AUB) associated with uterine fibroids (AUB-L) exhibits a molecular and cellular phenotype distinct from that observed in the endometrium from women with AUB associated with a primary endometrial disorder (AUB-E). Using clinical aetiology as a framework for comparative, menstrual cycle-staged transcriptomic and spatial analyses, we demonstrate that AUB-L is characterised by an altered global endometrial transcriptome and spatiotemporal immunolocalisation of sex steroid receptors compared to AUB-E. These findings demonstrate that a shared clinical symptom masks distinct underlying endometrial phenotypes.

These findings reposition the endometrium as an active mechanistic contributor, rather than a passive end-organ reacting to myometrial pathology. Previous investigations of AUB-L have focused primarily on fibroid biology for example, extracellular matrix remodelling, growth factor signalling, and altered vasculature architecture^27–29^. Conversely, studies of AUB-E historically emphasised endometrial angiogenesis, inflammation, haemostatic imbalance, and defective repair^28,30–32^. This direct *in vivo* molecular comparison of the endometrium, i.e., the tissue which bleeds, establishes that clinical causes of AUB, do not share a uniform pathway to bleeding.

To our knowledge, this is the first study to directly compare the *in vivo* endometrium across distinct clinical causes of AUB using well-defined patient cohorts. While historical investigations have grouped patients with AUB or compared them against presumptive “healthy” controls, defining “normal” menstruating endometrium remains highly challenging. Although researchers have used pregnancy as a biological marker of endometrial normality^21,33^, what constitutes physiological menstrual shedding and repair is poorly defined. By direct in vivo comparison of clinical causes of AUB, we separated the shared clinical symptom of bleeding from underlying aetiologies to interrogate endometrial biology. The distinct transcriptomic signature in AUB-L provides evidence of a “secondary endometrial disorder”^11^ where a structural cause alters the mucosal tissue at the cellular and molecular level. This aligns with evidence that the eutopic endometrium in symptomatic adenomyosis exhibits a transcriptomic signature entirely separate from the underlying structural lesion^34^.

Our findings also highlight the necessity of precise menstrual cycle staging. Menstrual cycle phase was the key determinant of molecular and cellular changes, aligning with physiological datasets related to the endometrium^13,14,21^. The importance of staging in patients with AUB has often been assumed but not rigorously reported. For example, a recent transcriptomic study compared “normal” endometrium to that of women with uterine fibroids and HMB, but pooled transcriptomic data across the entire secretory phase^35^. Given the transcriptome fluctuates dynamically across the secretory phase^21^, combining these data likely masks temporal, fibroid-related perturbations in the endometrium. We identified the mid-secretory stage as the critical window during which transcriptomic profiles and sex steroid receptor expression diverges, highlighting this specific menstrual cycle phase for future investigation.

A key finding of our present study is the differential spatiotemporal immunolocalisation of sex steroid receptors (PR, PR-B, and ERα) during the mid-secretory phase. While sex steroid receptor localisation in AUB-E aligned with physiological patterns previously described in functional cycling endometrium (including the expected loss of epithelial PR and PR-B)^13,14^, AUB-L was characterised by the persistence of glandular epithelial progesterone receptors alongside decreased stromal ERα expression. Crucially, these novel findings indicate that endometrial bleeding in AUB-L may be driven by aberrant spatial localisation of sex steroid hormone receptors, altering local tissue function, rather than a complete loss of hormone responsiveness.

This spatial variance may have therapeutic implications. “Progesterone resistance” in endometriotic lesions is defined by persistent PR expression^36^. The altered spatial PR and PR-B expression identified here in AUB-L, suggests a possible related mechanism that may explain why standard progestin therapies often fail in patients with AUB-L. If target sex steroid receptors are expressed in different endometrial cellular compartments depending on the underlying clinical cause of AUB, uniform, non-targeted progestins may not be universally effective across all patients presenting with AUB.

This observation builds upon in vitro models showing that endometrial epithelial organoids derived from women with uterine fibroids exhibit altered PR expression^37^. However, those organoids did not originate from patients presenting with the clinical symptom of AUB. Our *in vivo* findings address this translational gap, demonstrating that altered spatio-temporal receptor localisation may hold functional relevance for the abnormal endometrial bleeding phenotype.

Analysis of candidate genes, for example, *HOXA10* (a stromal marker of progesterone-regulated differentiation which rises in the secretory phase^14^) demonstrated secretory-phase downregulation in AUB-L, consistent with prior reports^38,39^. Together, these observations suggest progesterone signalling in AUB-L may be spatially modified rather than entirely absent, potentially reflecting disrupted stromal-epithelial crosstalk.

Unbiased transcriptomic analyses reinforce the aetiology-specific endometrial distinctions, identifying 81 DEGs when adjusted for menstrual cycle stage. Gene Ontology pathways related to platelet alpha granules and extracellular matrix were significantly upregulated in AUB-L, while ATP-dependent activity and chromatin remodelling pathways were downregulated. ATP-dependent pathways are central to cellular metabolic programming and directly underpin hormonally regulated decidualisation^40,41^. Label-free metabolic imaging has recently linked effective progesterone signalling to coordinated metabolic reprogramming^42^. Since decidualisation is essential for successful embryo implantation, and women with uterine fibroids have higher rates of implantation failure and early pregnancy loss^43^, an impaired decidual response, associated with altered PR localisation, disrupted *HOXA10* regulation^44^ and altered metabolic pathways, provides a potential biological mechanism linking abnormal bleeding with adverse fertility outcomes.

These novel data presented herein, raise the question as to whether endometrial metabolic pathways are differentially involved across different clinical causes of AUB *in vivo.* Although public databases often lack tissue-specific nuances for cycling endometrium^45^, our unbiased modelling suggests the global transcriptomic profile of AUB-L remains distinct from that of AUB-E.

Clinically, these findings challenge the standard empirical management of AUB, assumes that the bleeding endometrium responds to hormonal manipulation uniformly, regardless of the underlying clinical cause of AUB. The biological heterogeneity demonstrated herein may account for the variable efficacy of current, non-specific hormonal interventions; when these fail, approximately one-third of women ultimately undergo major surgery^46^. Hysterectomy remains the second most common surgical procedure among reproductive-aged women in the United States (>600,000 annually)^47^ and a leading intervention in the UK NHS (∼55,000 hysterectomies annually for HMB alone)^48^. This reliance on irreversible surgery highlights an important translational gap and an unmet need for precision medicine. Incorporating clinical phenotyping to target aetiology-specific endometrial mechanisms of AUB may improve patient outcomes and deliver wider societal value, with recent estimates suggesting every £1 invested in gynaecological care, yields over £11 to the wider economy^6^.

Study limitations include sample numbers constrained by strict inclusion criteria, and the fact that not all specimens underwent every analytical modality. Robust spatial and transcriptomic variations were nevertheless observed. Future prospective clinical trials evaluating *in vivo* progestin efficacy alongside spatial receptor profiling are required to associate these molecular profiles with clinical outcomes. Additionally, spatial-transcriptomics and single-cell sequencing are needed to further delineate cell-specific contributions^3^.

In summary, this study provides in vivo evidence that clinically distinct causes of AUB (AUB-L and AUB-E) are associated with biologically distinct endometrial profiles. Despite presenting with the identical clinical symptom of abnormal bleeding, transcriptomic differences and altered spatial localisation of sex steroid receptors indicate that AUB is not a single clinical entity. These findings support a paradigm shift in gynaecological care from empirical, uniform treatment toward precision phenotyping and therapeutics driven by the underlying cause of AUB, i.e. a precision medicine-based approach.

## Acknowledgements

This work was supported with funding from the UK Charity Wellbeing of Women (RTF902), BBSRC-NC3R ageing project grant BB/S002995/1; Wellcome Trust (225021/Z/22/Z); MRC Centre for Reproductive Health (MRC CRH) Centre grants: G1002033, MR/N022556/1; MRC research grants G0000066, G0500047, G0600048, and MR/J003611/1.

We are grateful for the valuable contributions to this study from our clinical research team: Sharon MacPherson, Catherine Murray, Moira Nicol, Alison Murray; staff from the University of Edinburgh facilities: Edinburgh Genomics and SURF histology for their support, and the many patients who have so generously donated their tissues for clinical research.

## Data Availability

Data will be made available with publication.

