## Supplemental Materials for "Abnormal uterine bleeding (AUB) is not a single clinical entity: different clinical causes are associated with distinct cellular and molecular endometrial characteristics"

**Supplementary Material: Table of Contents**

| | Page Numbers |
|---|---|
| Supplementary Table 1 | 2-5 |
| Supplementary Table 2 | 6 |
| Supplementary Table 3 | 6 |
| Supplementary Table 4 | 7-8 |
| Supplementary Table 5 | 9 |

**Supplementary Table 1:** Characteristics for patients from whom endometrial samples were used within this study. 73 endometrial samples, were categorised according to i) the associated cause of abnormal uterine bleeding (AUB) determined through clinical history and investigations (i.e., AUB-L refers to the presence of fibroids, AUB-E refers to primary endometrial disorder), ii) stage of menstrual cycle determined by combined assessment of histological review of the endometrial tissue sample, serum levels of oestradiol ($E_2$) and progesterone ($P_4$) taken at the time of the endometrial sampling and menstrual cycle day and menstrual cycle length were recorded from patient recall, referred to as the "Edinburgh Classification". The source of the tissue sample could either be an endometrial biopsy (B) or hysterectomy (H). All samples were interrogated using RT-qPCR. A cohort of samples were studied using immunohistochemistry (IHC) and a different cohort were studied using bulk RNA sequencing. If endometrial samples underwent bulk RNA sequencing, their menstrual cycle timepoint (%), as determined by the Endest[21] algorithm, has been provided. If cells within the table contain a "-", this means the data are missing from the details collected during recruitment.

| | **Cause of AUB** | **Menstrual cycle stage as per "Edinburgh Classification"** | **Source of biopsy** | **Age Range (5 yr group)** | **BMI** | **Parity** | **Cycle Day** | **Cycle Length** | **Serum $E_2$ (pmol/l)** | **Serum $P_4$ (nmol/l)** | **IHC** | **Bulk RNA seq** | **Endest[21] menstrual cycle timepoint (%)** |
|---|---|---|---|---|---|---|---|---|---|---|---|---|---|
| 1 | AUB-E | Proliferative | H | 35-39 | 33 | 5 | 10 | | 275 | <3 | | | |
| 2 | AUB-E | Proliferative | H | 40-44 | - | 2 | 14 | 30 | 921 | 8 | | | |
| 3 | AUB-E | Proliferative | B | 30-34 | 43 | 2 | 15 | 35 | 349 | 1 | | | |
| 4 | AUB-E | Proliferative | H | 35-39 | 35 | 2 | 8 | 28 | 215 | <3 | | yes | 23 |
| 5 | AUB-E | Proliferative | H | 45-49 | 22 | 2 | 8 | 24 | 7 | 2 | yes | | |
| 6 | AUB-E | Proliferative | B | 40-44 | 29 | 2 | 9 | 29 | 281 | <3 | | yes | 26 |
| 7 | AUB-E | Proliferative | B | 35-39 | 21 | - | 11 | 27 | 1359 | 1 | | | |
| 8 | AUB-E | Proliferative | B | 30-34 | 20 | 1 | 6 | 25 | 120 | 1 | | | |
| 9 | AUB-E | Proliferative | B | 35-39 | - | 3 | 9 | 30 | 97 | 0 | | | |
| 10 | AUB-E | Proliferative | B | 35-39 | 38 | 1 | 13 | 25 | 4 | 1 | | yes | 28 |
| 11 | AUB-E | Proliferative | H | 25-29 | 23 | 3 | 18 | 28 | 320 | 1 | | | |
| 12 | AUB-E | Proliferative | B | 35-39 | 47 | 3 | 16 | 24 | 495 | 3 | | | |
| 13 | AUB-E | Proliferative | B | 35-39 | 21 | 0 | 15 | 31 | 314 | 7 | | | |
| 14 | AUB-E | Proliferative | B | 40-44 | 31 | 2 | 13 | 30 | 607 | 1 | yes | | |
| 15 | AUB-E | Proliferative | B | 40-44 | - | 0 | 10 | 32 | 168 | 2 | | | |
| 16 | AUB-E | Proliferative | B | 40-44 | 33 | 1 | 11 | 31 | 647 | 0 | | yes | 28 |
| 17 | AUB-E | Proliferative | B | 40-44 | 23 | 3 | 15 | 25 | 398 | 4 | | yes | 44 |
| 18 | AUB-E | Proliferative | B | 40-44 | 21 | 2 | 11 | 25 | 271 | 1 | | | |

| | | | | | | | | | | | | | |
|---|---|---|---|---|---|---|---|---|---|---|---|---|---|
| 19 | AUB-E | Proliferative | B | 40-44 | 21 | 1 | 10 | 28 | 723 | 0 | yes | | |
| 20 | AUB-E | Proliferative | B | 45-49 | 40 | 2 | 17 | 31 | 169 | 2 | | | |
| 21 | AUB-E | Proliferative | B | 45-49 | 26 | 2 | | | 196 | 0 | yes | yes | 21 |
| 22 | AUB-E | Proliferative | B | 50-54 | 31 | 3 | 14 | 28 | 1461 | 2 | yes | | |
| 1 | AUB-L | Proliferative | H | 40-44 | - | 0 | | 28 | 67 | 4 | | yes | 11 |
| 2 | AUB-L | Proliferative | H | 40-44 | - | 2 | 8 | 32 | 971 | 4 | | | |
| 3 | AUB-L | Proliferative | B | 40-44 | 19 | 2 | 7 | 28 | 491 | <3 | yes | | |
| 4 | AUB-L | Proliferative | B | 45-49 | 43 | 3 | 11 | 28 | 879 | 1 | | | |
| 5 | AUB-L | Proliferative | B | 50-54 | 26 | 1 | 12 | 28 | 96 | 4 | | | |
| 6 | AUB-L | Proliferative | B | 45-49 | - | 2 | 8 | 28 | 185 | 1 | yes | | |
| 7 | AUB-L | Proliferative | B | 45-49 | 32 | 1 | 17 | 30 | 221 | 6 | | yes | 47 |
| 8 | AUB-L | Proliferative | B | 40-44 | 21 | 2 | 8 | 32 | 672 | <3 | | | |
| 9 | AUB-L | Proliferative | B | 50-54 | 27 | 1 | 14 | 28 | 312 | 1 | yes | | |
| 10 | AUB-L | Proliferative | B | 35-39 | 41 | 2 | 16 | 28 | 333 | 7 | yes | yes | 52 |
| 11 | AUB-L | Proliferative | B | 40-44 | 31 | 0 | 15 | 25 | 878 | 1 | yes | yes | 27 |
| 12 | AUB-L | Proliferative | B | 40-44 | 37 | 0 | 12 | 30 | 501 | 0 | yes | | |
| 13 | AUB-L | Proliferative | B | 45-49 | 22 | 2 | | | 426 | 2 | | yes | 40 |
| 14 | AUB-L | Proliferative | H | 45-49 | - | 3 | 9 | 28 | 1685 | 4 | | yes | 20 |
| 1 | AUB-E | Early Secretory | B | 30-34 | 22 | 2 | 16 | 28 | 560 | 41 | | yes | 65 |
| 2 | AUB-E | Early Secretory | H | 30-34 | - | 2 | 22 | 29 | 406 | 59 | | yes | 67 |
| 1 | AUB-L | Early Secretory | H | 45-49 | - | 3 | 23 | 28 | 682 | 15 | | | |
| 2 | AUB-L | Early Secretory | B | 45-49 | 21 | 1 | | 28 | 884 | 52 | | yes | 65 |
| 3 | AUB-L | Early Secretory | B | 40-44 | 30 | 3 | 21 | 25 | 619 | 62 | | yes | 67 |
| 4 | AUB-L | Early Secretory | H | 35-39 | - | 3 | 16 | 28 | 848 | 93 | | yes | 63 |
| 5 | AUB-L | Early Secretory | B | 40-44 | 38 | 0 | 19 | 29 | 620 | 32 | | yes | 65 |
| 6 | AUB-L | Early Secretory | H | 40-44 | - | 2 | 18 | 28 | 401 | 72 | | | |

| | | | | | | | | | | | | | |
|---|---|---|---|---|---|---|---|---|---|---|---|---|---|
| 7 | AUB-L | Early Secretory | B | 40-44 | 22 | 2 | 18 | 31 | 458 | 22 | | | |
| 8 | AUB-L | Early Secretory | B | 40-44 | 26 | 0 | 16 | 30 | 218 | 30 | | | |
| 9 | AUB-L | Early Secretory | B | 40-44 | 20 | 3 | 9 | 24 | 154 | 42 | | | |
| 10 | AUB-L | Early Secretory | B | 45-49 | 30 | 4 | | 30 | 354 | 42 | | yes | 61 |
| 11 | AUB-L | Early Secretory | B | 50-54 | 25 | 5 | 23 | | 218 | 25 | | | |
| 1 | AUB-E | Early to Mid-Secretory | B | 25-29 | - | 4 | 22 | 30 | 395 | 88 | | | |
| 2 | AUB-E | Early to Mid-Secretory | B | 30-34 | - | 2 | 28 | | 450 | 45 | | yes | 70 |
| 3 | AUB-E | Early to Mid-Secretory | B | 35-39 | 33 | 1 | 15 | 21 | 85 | 23 | | | |
| 4 | AUB-E | Early to Mid-Secretory | B | 45-49 | 43 | 2 | 15 | | 242 | 47 | | yes | 71 |
| 5 | AUB-E | Early to Mid-Secretory | B | 40-44 | 32 | 2 | 18 | 28 | 469 | 14 | | yes | 75 |
| 6 | AUB-E | Early to Mid-Secretory | H | 40-44 | - | 2 | 18 | 28 | 502 | 43 | | yes | 67 |
| 1 | AUB-L | Early to Mid-Secretory | B | 45-49 | 24 | 2 | 14 | 28 | 321 | 22 | | yes | 60 |
| 1 | AUB-E | Mid-secretory | B | 45-49 | 30 | 0 | 23 | 29 | | - | | yes | 72 |
| 2 | AUB-E | Mid-secretory | B | 40-44 | 27 | 0 | | 28 | 41 | 347 | yes | yes | 76 |
| 3 | AUB-E | Mid-secretory | B | 40-44 | 30 | 1 | 18 | | 392 | 22 | yes | | |
| 4 | AUB-E | Mid-secretory | B | 35-39 | 20 | 3 | 24 | 28 | 326 | 26 | yes | yes | 80 |
| 5 | AUB-E | Mid-secretory | B | 35-39 | 24 | 0 | 29 | 28 | 353 | 43 | | yes | 80 |
| 6 | AUB-E | Mid-secretory | B | 50-54 | 24 | 0 | 24 | 35 | 271 | 21 | | yes | 78 |
| 7 | AUB-E | Mid-secretory | H | 40-44 | - | 2 | 23 | 28 | 331 | 84 | | | |
| 8 | AUB-E | Mid-secretory | B | 35-39 | - | 2 | 26 | 32 | 627 | 94 | | yes | 84 |
| 9 | AUB-E | Mid-secretory | B | 40-44 | - | 1 | 20 | | 456 | 40 | yes | | |
| 10 | AUB-E | Mid-secretory | B | 35-39 | 23 | 1 | 25 | 30 | 145 | 24 | | | |
| 11 | AUB-E | Mid-secretory | B | 40-44 | 25 | 3 | 28 | 30 | 716 | 36 | | | |
| 1 | AUB-L | Mid-secretory | H | 40-44 | 22 | 1 | 24 | 22 | 396 | 36 | yes | yes | 84 |
| 2 | AUB-L | Mid-secretory | B | 45-49 | 43 | 5 | 14 | 28 | 361 | 29 | | yes | 76 |

| | | | | | | | | | | | | | |
|---|---|---|---|---|---|---|---|---|---|---|---|---|---|
| 3 | AUB-L | Mid-secretory | B | 50-54 | 27 | 2 | 15 | 28 | 127 | 32 | | yes | 79 |
| 4 | AUB-L | Mid-secretory | B | 40-44 | 20 | 0 | 34 | 35 | 1139 | 16 | yes | yes | 59 |
| 5 | AUB-L | Mid-secretory | B | 40-44 | - | 2 | 23 | 31 | 620 | 23 | yes | yes | 73 |
| 6 | AUB-L | Mid-secretory | B | 45-49 | 27 | 2 | | 27 | 487 | 47 | | | |

**Supplementary Table 2:** Primers and probes for RT-qPCR (Universal Probe Library, Roche Applied Science, USA) for ATP synthase F1 subunit beta (*ATP5B*), Succinate dehydrogenase complex flavoprotein subunit A (*SDHA*), progesterone receptor (*PGR*), progesterone receptor B (*PRB2*), oestrogen Receptor (*ESR1*), Interleukin 15 (*IL15*), Heart- and Neural Crest Derivatives-Expressed Protein 2 (*HAND2*), Forkhead Box O1 (*FOXO1*), Homeobox A10 (*HOXA10*), 17β-Hydroxysteroid dehydrogenase 2 (*HSD17B2*).

| Gene of Interest | Forward Primer | Reverse Primer | Roche Probe (UPL) |
|---|---|---|---|
| ***Housekeeping genes*** | | | |
| *ATP5F1B* | Agaggtcccatcaaaaccaa | tcctgctcaacactcatttcc | 50 |
| *SDHA* | Tccactacatgacggagcag | Ccatcttcagttctgctaaacg | 70 |
| ***Steroid Receptors*** | | | |
| *PGR* | Tttaagagggcaatggaagg | cggattttatcaacgatgcag | 11 |
| *ESR1* | Tgggtgattgccaagag | agatgttccatgccctt | 52 |
| ***Progesterone Dependent Genes*** | | | |
| *IL15* | Cagatagccagcccatacaag | ggctatggcaaggggttt | 46 |
| *HAND2* | Tcaagaagaccgacgtgaaa | Gttgctgctcactgtgcttt | 35 |
| *FOXO1* | Aagggtgacagcaacagctc | ttccttcattctgcacacga | 11 |
| *HOXA10* | Ccttccgagagcagcaaa | ttggctgcgttttcacct | 61 |
| ***Steroid Hormone Metabolising Enzyme*** | | | |
| *HSD17B2* | Agggaggctggtgaatgtc | cgcctttgatgagccataag | 52 |

**Supplementary Table 3:** Primary antibodies investigated using automated immunohistochemistry (LEICA BOND)

| Antibody | Supplier Reference | Antibody Type | Antibody Concentration | Antibody Dilution (in NHS) | Negative Control | BOND Refine Protocol | BOND Epitope Retrieval Buffer |
|---|---|---|---|---|---|---|---|
| Progesterone Receptor | Dako M3569 | Monoclonal Mouse | 80.8mg/l | 1 in 200 | Mouse 1gG | 60'15'15' (mouse) | ER1 |
| Progesterone Receptor B | Cell Signalling 3157S | Monoclonal Rabbit | 243mcg/ml | 1 in 800 | Rabbit 1gG | 60'15' no pre-polymer (rabbit) | ER1 |
| Oestrogen Receptor alpha | Dako M3643 | Monoclonal Rabbit | 149mg/l | 1 in 200 | Rabbit IgG | 60'15' no pre-polymer (rabbit) | ER1 |

**<u>Supplementary Table 4</u>**: Significant differentially expressed genes (DEGs) identified when comparing whole transcriptome RNA sequencing from 17 endometrial samples from women with AUB associated with uterine fibroids (AUB-L) versus 18 endometrial samples from women with AUB associated with a primary endometrial disorder (AUB-E). Proliferative phase comparison did not reveal any significant DEGs).

| ENSEMBL | Gene name | logFC | AveExpr | t | P.Value | adj.P.Val | B |
|---|---|---|---|---|---|---|---|
| ENSG00000183682 | *BMP8A* | 1.06 | 1.34 | 6.637 | 1.97E-07 | 0.003211 | 6.76 |
| ENSG00000103742 | *IGDCC4* | 1.478 | 0.07675 | 5.873 | 1.72E-06 | 0.01027 | 4.195 |
| ENSG00000154262 | *ABCA6* | -1.118 | 1.147 | -5.783 | 2.22E-06 | 0.01027 | 4.387 |
| ENSG00000176435 | *CLEC14A* | 0.7358 | 3.78 | 5.74 | 2.51E-06 | 0.01027 | 4.753 |
| ENSG00000256968 | *Not assigned* | 1.919 | -0.1349 | 5.618 | 3.57E-06 | 0.01166 | 3.681 |
| ENSG00000233270 | *Not assigned* | 1.45 | 0.1193 | 5.514 | 4.82E-06 | 0.01311 | 3.341 |
| ENSG00000094841 | *UPRT* | 0.6593 | 2.894 | 5.31 | 8.66E-06 | 0.01783 | 3.56 |
| ENSG00000138131 | *LOXL4* | 1.046 | 1.96 | 5.307 | 8.73E-06 | 0.01783 | 3.459 |
| ENSG00000148180 | *GSN* | 0.479 | 7.892 | 5.172 | 1.29E-05 | 0.02336 | 3.208 |
| ENSG00000112303 | *VNN2* | 1.11 | 0.272 | 5.135 | 1.43E-05 | 0.02336 | 2.768 |
| ENSG00000168994 | *PXDC1* | 0.6849 | 3.1 | 5.043 | 1.87E-05 | 0.02634 | 2.877 |
| ENSG00000162738 | *VANGL2* | 0.816 | 2.055 | 5.007 | 2.07E-05 | 0.02634 | 2.767 |
| ENSG00000108684 | *ASIC2* | 1.174 | 0.3177 | 5.002 | 2.10E-05 | 0.02634 | 2.707 |
| ENSG00000153485 | *LYSET* | 0.9115 | 3.303 | 4.941 | 2.50E-05 | 0.02757 | 2.606 |
| ENSG00000254585 | *MAGEL2* | 1.472 | 0.03228 | 4.904 | 2.78E-05 | 0.02757 | 2.242 |
| ENSG00000165219 | *GAPVD1* | -0.5935 | 5.493 | -4.899 | 2.82E-05 | 0.02757 | 2.485 |
| ENSG00000225616 | *RPL27P4* | 1.865 | 0.7952 | 4.893 | 2.87E-05 | 0.02757 | 2.162 |
| ENSG00000172936 | *MYD88* | 0.5212 | 4.259 | 4.862 | 3.14E-05 | 0.02849 | 2.39 |
| ENSG00000271447 | *MMP28* | 1.019 | -0.7031 | 4.7 | 5.00E-05 | 0.03979 | 1.535 |
| ENSG00000165288 | *BRWD3* | -0.5783 | 4.575 | -4.685 | 5.21E-05 | 0.03979 | 1.925 |
| ENSG00000175130 | *MARCKSL1* | 0.6376 | 6.068 | 4.644 | 5.86E-05 | 0.03979 | 1.777 |
| ENSG00000213553 | *RPLP0P6* | 0.8302 | 0.4817 | 4.641 | 5.92E-05 | 0.03979 | 1.582 |
| ENSG00000150687 | *PRSS23* | 0.6903 | 7.318 | 4.608 | 6.50E-05 | 0.03979 | 1.681 |
| ENSG00000202538 | *RNU4-2* | -1.082 | 0.8622 | -4.578 | 7.08E-05 | 0.03979 | 1.431 |
| ENSG00000240509 | *RPL34P18* | 0.9364 | 2.853 | 4.575 | 7.14E-05 | 0.03979 | 1.64 |
| ENSG00000100403 | *ZC3H7B* | 0.4473 | 5.782 | 4.57 | 7.25E-05 | 0.03979 | 1.577 |
| ENSG00000271601 | *LIX1L* | 0.6778 | 4.689 | 4.565 | 7.33E-05 | 0.03979 | 1.576 |
| ENSG00000196584 | *XRCC2* | -0.8184 | 1.331 | -4.558 | 7.48E-05 | 0.03979 | 1.394 |
| ENSG00000050165 | *DKK3* | 0.6117 | 4.063 | 4.555 | 7.55E-05 | 0.03979 | 1.56 |
| ENSG00000087076 | *HSD17B14* | 0.705 | 1.563 | 4.549 | 7.68E-05 | 0.03979 | 1.556 |
| ENSG00000198515 | *CNGA1* | 1.089 | 0.6827 | 4.547 | 7.74E-05 | 0.03979 | 1.436 |
| ENSG00000138459 | *SLC35A5* | 0.8046 | 3.697 | 4.534 | 8.03E-05 | 0.03979 | 1.519 |
| ENSG00000119699 | *TGFB3* | 0.6235 | 2.769 | 4.533 | 8.04E-05 | 0.03979 | 1.526 |
| ENSG00000065320 | *NTN1* | 0.5846 | 3.418 | 4.515 | 8.47E-05 | 0.04067 | 1.472 |
| ENSG00000138031 | *ADCY3* | 0.6321 | 4.787 | 4.505 | 8.71E-05 | 0.04067 | 1.421 |
| ENSG00000157106 | *SMG1* | -0.5024 | 5.079 | -4.486 | 9.19E-05 | 0.04072 | 1.389 |
| ENSG00000069943 | *PIGB* | 0.575 | 2.872 | 4.477 | 9.45E-05 | 0.04072 | 1.385 |
| ENSG00000135916 | *ITM2C* | 0.5847 | 4.532 | 4.474 | 9.50E-05 | 0.04072 | 1.334 |
| ENSG00000171051 | *FPR1* | 1.25 | -0.1597 | 4.466 | 9.72E-05 | 0.04072 | 0.9518 |
| ENSG00000128284 | *APOL3* | 0.5453 | 2.868 | 4.429 | 0.0001081 | 0.04257 | 1.255 |

| | | | | | | | |
|---|---|---|---|---|---|---|---|
| ENSG00000145936 | *KCNMB1* | 1.26 | 0.1217 | 4.428 | 0.0001084 | 0.04257 | 1.074 |
| ENSG00000155966 | *AFF2* | -1.181 | 1.739 | -4.42 | 0.0001111 | 0.04257 | 1.14 |
| ENSG00000100325 | *ASCC2* | 0.4056 | 5.551 | 4.416 | 0.0001121 | 0.04257 | 1.171 |
| ENSG00000034533 | *ASTE1* | -0.5437 | 2.707 | -4.405 | 0.0001159 | 0.04303 | 1.185 |
| ENSG00000101000 | *PROCR* | 0.7739 | 3.979 | 4.391 | 0.0001205 | 0.04353 | 1.143 |
| ENSG00000104517 | *UBR5* | -0.452 | 5.974 | -4.383 | 0.0001233 | 0.04353 | 1.085 |
| ENSG00000198276 | *UCKL1* | 0.6743 | 3.493 | 4.374 | 0.0001265 | 0.04353 | 1.117 |
| ENSG00000111676 | *ATN1* | 0.611 | 5.767 | 4.37 | 0.0001279 | 0.04353 | 1.042 |
| ENSG00000102760 | *RGCC* | 1.174 | 3.116 | 4.362 | 0.0001307 | 0.04356 | 1.079 |
| ENSG00000161642 | *ZNF385A* | 0.8009 | 1.828 | 4.345 | 0.0001372 | 0.04483 | 0.9884 |
| ENSG00000167565 | *SERTAD3* | 0.8726 | 2.091 | 4.334 | 0.0001417 | 0.0454 | 0.9977 |
| ENSG00000115317 | *HTRA2* | 0.4289 | 4.147 | 4.309 | 0.000152 | 0.04768 | 0.9194 |
| ENSG00000086205 | *FOLH1* | 0.8223 | 1.563 | 4.303 | 0.0001547 | 0.04768 | 0.8894 |
| ENSG00000065154 | *OAT* | 0.6323 | 5.295 | 4.275 | 0.0001674 | 0.0489 | 0.8042 |
| ENSG00000204387 | *SNHG32* | 1.431 | 0.1977 | 4.271 | 0.0001692 | 0.0489 | 0.44 |
| ENSG00000270179 | *Not assigned* | 0.9699 | 0.4503 | 4.258 | 0.0001753 | 0.0489 | 0.5621 |
| ENSG00000124785 | *NRN1* | 0.7724 | 0.6958 | 4.258 | 0.0001757 | 0.0489 | 0.753 |
| ENSG00000198894 | *CIPC* | 0.728 | 4.131 | 4.257 | 0.0001759 | 0.0489 | 0.7756 |
| ENSG00000211751 | *TRBC1* | 0.581 | 2.296 | 4.246 | 0.0001813 | 0.0489 | 0.7936 |
| ENSG00000198482 | *ZNF808* | -0.5702 | 3.464 | -4.242 | 0.0001835 | 0.0489 | 0.7845 |
| ENSG00000132849 | *PATJ* | -0.5817 | 5.213 | -4.241 | 0.0001839 | 0.0489 | 0.7678 |
| ENSG00000144554 | *FANCD2* | -0.5997 | 3.783 | -4.238 | 0.0001857 | 0.0489 | 0.7702 |
| ENSG00000134058 | *CDK7* | 0.4823 | 3.809 | 4.232 | 0.0001886 | 0.0489 | 0.7336 |
| ENSG00000168487 | *BMP1* | 0.4878 | 4.432 | 4.208 | 0.0002022 | 0.04988 | 0.6303 |
| ENSG00000183918 | *SH2D1A* | 1.201 | 0.009397 | 4.198 | 0.0002077 | 0.04988 | 0.1788 |
| ENSG00000126883 | *NUP214* | -0.3252 | 5.553 | -4.197 | 0.0002082 | 0.04988 | 0.5928 |
| ENSG00000146707 | *POMZP3* | -1.214 | 0.565 | -4.195 | 0.0002094 | 0.04988 | 0.4059 |
| ENSG00000275342 | *PRAG1* | 0.6044 | 2.172 | 4.194 | 0.00021 | 0.04988 | 0.6571 |
| ENSG00000138802 | *SEC24B* | -0.3587 | 4.227 | -4.174 | 0.0002226 | 0.04988 | 0.5796 |
| ENSG00000047056 | *WDR37* | 0.4548 | 2.555 | 4.173 | 0.0002232 | 0.04988 | 0.6079 |
| ENSG00000257181 | *Not assigned* | -1.697 | 0.337 | -4.171 | 0.0002242 | 0.04988 | 0.08666 |
| ENSG00000133250 | *ZNF414* | 0.4779 | 4.2 | 4.171 | 0.0002243 | 0.04988 | 0.5517 |
| ENSG00000224114 | *RPS14P1* | 2.639 | -0.04735 | 4.169 | 0.0002252 | 0.04988 | 0.4227 |
| ENSG00000138796 | *HADH* | 0.4952 | 5.623 | 4.162 | 0.00023 | 0.04988 | 0.4954 |
| ENSG00000198794 | *SCAMP5* | 0.7545 | 2.272 | 4.152 | 0.0002368 | 0.04988 | 0.5455 |
| ENSG00000018625 | *ATP1A2* | 0.9615 | -0.2852 | 4.145 | 0.0002409 | 0.04988 | 0.5364 |
| ENSG00000143226 | *FCGR2A* | 0.6822 | 0.9034 | 4.145 | 0.0002411 | 0.04988 | 0.5006 |
| ENSG00000100842 | *EFS* | 0.474 | 4.419 | 4.141 | 0.000244 | 0.04988 | 0.4571 |
| ENSG00000185305 | *ARL15* | 0.4663 | 3.798 | 4.139 | 0.0002455 | 0.04988 | 0.477 |
| ENSG00000172432 | *GTPBP2* | 0.9367 | 3.244 | 4.136 | 0.0002471 | 0.04988 | 0.5028 |
| ENSG00000176915 | *ANKLE2* | -0.4396 | 5.13 | -4.136 | 0.0002474 | 0.04988 | 0.453 |

**Supplementary Table 5**: Pathways identified to be significantly over-represented when conducting a gene ontology enrichment analysis using the 64 significant upregulated differentially expressed genes (DEGs) and the 17 significant downregulated DEGs. Pathways have been divided as those which were identified to be upregulated or downregulated in the endometrium from women with AUB associated with uterine fibroids (AUB-L) when compared to endometrium from women with AUB associated a primary endometrial disorder (AUB-E).

| **Pathways upregulated in endometrium from women with AUB-L** | | | | | | | | |
|---|---|---|---|---|---|---|---|---|
| **ID** | **Description** | **GeneRatio** | **BgRatio** | **pvalue** | **p.adjust** | **qvalue** | **geneID** | **Count** |
| GO:0031093 | platelet alpha granule lumen | 7/266 | 48/13228 | 4.47E-05 | 0.01271 | 0.0125 | *TGFB3/ GTPBP2/ SPARC/ IGF2/ FERMT3/ MMRN1/ F8* | 7 |
| GO:0031091 | platelet alpha granule | 8/266 | 69/13228 | 6.97E-05 | 0.01271 | 0.0125 | *TGFB3/ GTPBP2/ SPARC/ SNCA/ IGF2/ FERMT3/ MMRN1/ F8* | 8 |
| GO:0031012 | extracellular matrix | 19/266 | 390/13228 | 0.000347 | 0.03268 | 0.03213 | *CLEC14A/ LOXL4/ MMP28/ TGFB3/ NTN1/ SPARC/ AEBP1/ FBLN2/ PODN/ LRRTM1/ MFAP4/ COL8A2/ EFEMP1/ WNT7A/ ANXA5/ ADAMTSL5/ WNT5B/ MMRN1/ SNED1* | 19 |
| GO:0030312 | external encapsulating structure | 19/266 | 391/13228 | 0.000358 | 0.03268 | 0.03213 | *CLEC14A/ LOXL4/ MMP28/ TGFB3/ NTN1/ SPARC/ AEBP1/ FBLN2/ PODN/ LRRTM1/ MFAP4/ COL8A2/ EFEMP1/ WNT7A/ ANXA5/ ADAMTSL5/ WNT5B/ MMRN1/ SNED1* | 19 |
| **Pathways downregulated in endometrium from women with AUB_L** | | | | | | | | |
| **ID** | **Description** | **GeneRatio** | **BgRatio** | **pvalue** | **p.adjust** | **qvalue** | **geneID** | **Count** |
| GO:0140657 | ATP-dependent activity | 16/101 | 486/13028 | 9.43E-07 | 0.000258 | 0.000251 | *ABCA6/XRCC2/DHX40/TDRD12/ATP2B1/CHD7/CHD8/KIF1B/NAV2/BRIP1/CECR2/SMCHD1/ERCC6/HELZ/ACSL3/CDC6* | 16 |
| GO:0008094 | ATP-dependent activity, acting on DNA | 7/101 | 109/13028 | 2.13E-05 | 0.001968 | 0.001913 | *XRCC2/CHD7/CHD8/NAV2/BRIP1/CECR2/ERCC6* | 7 |
| GO:0016887 | ATP hydrolysis activity | 12/101 | 360/13028 | 2.15E-05 | 0.001968 | 0.001913 | *ABCA6/DHX40/TDRD12/ATP2B1/CHD7/CHD8/KIF1B/NAV2/BRIP1/SMCHD1/ERCC6/CDC6* | 12 |
| GO:0004386 | helicase activity | 7/101 | 139/13028 | 0.000101 | 0.006916 | 0.006722 | *DHX40/TDRD12/CHD7/CHD8/NAV2/BRIP1/HELZ* | 7 |
| GO:0140658 | ATP-dependent chromatin remodeler activity | 4/101 | 34/13028 | 0.000132 | 0.007233 | 0.00703 | *CHD7/CHD8/CECR2/ERCC6* | 4 |
